# Severe underestimation of COVID-19 case numbers: effect of epidemic growth rate and test restrictions

**DOI:** 10.1101/2020.04.13.20064220

**Authors:** Peter Richterich

## Abstract

To understand the scope and development of the COVID-19 pandemic, knowledge of the number of infected persons is essential. Often, the number of “confirmed cases”, which is based on positive RT-PCR test results, is regarded as a reasonable indicator. However, limited COVID-19 test capacities in many countries are restricting the amount of testing that can be done. This can lead to the implementation of testing policies that restrict access to COVID-19 tests, and to testing backlogs and delays. As a result, confirmed case numbers can be significantly lower than the actual number of infections, especially during rapid growth phases of the epidemic.

This study examines the quantitative relation between infections and reported confirmed case numbers for two different testing strategies, “limited” and “inclusive” testing, in relation to the growth rate of the epidemic. The results indicate that confirmed case numbers understate the actual number of infections substantially; during rapid growth phases where the daily growth rate can reach or exceed 30%, as has been seen in many countries, the confirmed case numbers under-report actual infections by up to 50 to 100-fold.

## Introduction

The COVID-19 pandemic is a major global health threat, with more than 1.9 million confirmed cases and 118,854 deaths as of April 13, 2020. Understanding the current state of the epidemic in a country or region requires knowing the actual number of infections, which relies on testing possibly infected persons for the SARS-CoV-2 virus.

Different countries have adopted very different COVID-19 testing strategies, which is often dictated by a combination of perceived risk and resource constraints. Several countries that had been hit hard by the SARS and MERS epidemics implemented comprehensive testing strategies that aimed to test every infected person and every person who was exposed to an infected person.^1–3^ In many other countries, more limited testing strategies are used, where testing is restricted to a subset of the infected. Typical criteria required to qualify for testing are severe symptoms; prior contact with a confirmed COVID-19 case; belonging to a “high-risk” group; or a combination of these and other restrictions. In countries that have limited testing, access to tests often requires a referral by a medical doctor, and anecdotal evidence suggests that tests are sometimes denied even to persons with a high likelihood of being infected.

When only a small subset of infected persons is tested, the results is a higher “raw case fatality ratio (CFR)”, defined as the number of COVID-19 fatalities divided confirmed cases. Observed CFR ratios have varied substantially between countries and over time. For example, on March 25, 2020, Germany had a raw CFR of 0.55%, while Italy had an 18-fold higher CFR of 10.1%. On April 10, 2020, Germany’s raw CFR had increased fourfold, to 2.26%

A number of studies have concluded that different test rates are the primary cause of the observed differences in fatality rates. Such analyses are often based on knowledge and assumptions about “real” CFR rates and observed death rates. Multiple studies have shown that the reported numbers of confirmed cases may underestimate the true number of cases by a factor of 10 or even more.^4–6^ While such studies can provide useful insights, they often cannot explain the observed change in the CFR during an epidemic.

Contributing to the under-reporting of COVID-19 cases is a combination of two factors: (a) the rapid growth of the epidemics, and (b) the incubation period of 5 to 6 days. In many countries, the observed doubling times for the COVID-19 epidemic has been 3 days or even shorter for extended periods. In such rapid growth phases, the number of infected persons doubles *twice* in the time between infection and symptom onset. Even after the onset of symptoms, additional delays can occur before getting a test, and again to have the test results included in the officially reported numbers; these delays can allow for another doubling of infections. To give an example, let us assume an incubation period of 6 days, and another 3-day delay to get the test. By the time the test results become available after 9 days, the size of the epidemic has doubled three times in succession, for an overall 8-fold growth; 7 of the 8 infections occurred after the initial infection, and are either still in the incubation period, waiting to get tested, or waiting for test results. In other words, confirmed cases would under-report existing infections by a factor of 8.

The exact magnitude of the “under-reporting factor” depends on the actual growth of the epidemic, testing and reporting delays, and testing policy. In this study, I describe the results of modeling studies that estimate the magnitude of the under-reporting factor.

## Methods

Data about COVID-19 cases and deaths were downloaded from the Johns Hopkins University dataset at https://github.com/CSSEGISandData/COVID-19/.

### SIRD model

We developed an iterative model similar to standard Susceptible/ Exposed/ Infectious/ Recovered/ Dead (SEIRD) models, with several modifications aimed to model the biology of viral infections in a way that is straightforward to implement. At the beginning of the simulation, the size of the susceptible group is set to the size of the population being modeled (i.e. 327 million for the US). The simulation starts with a small number of individuals (for instance four) in the “infected” category.

Rather than using an explicit separation of non-infectious “Exposed” individuals and a separate “Infectious” group, we track infections by “infection age”, i.e. the number of days since an individual was exposed. Individuals with the same “infection age” are grouped together, and tracked using a single counter. In total, the infected population is tracked for 30 days, and then moved to the “Recovered” group.

To reflect the time-dependent infectiousness of individuals^7^, infectivity is modeled using a per-day infectivity array that reflects current estimates of infectivity during and after the incubation period. We used a scaled gamma distribution, where the value for each day represents the cumulative probability of infection for the previous 24 hours, and a fixed offset for a non-infectious period immediately after infection. Note that the scale factor used is equivalent to the initial reproduction rate R^0^. Function parameters are given in Table 1. The resulting unscaled per-day infectivity factors are shown in Figure 1.

**Table 1:**
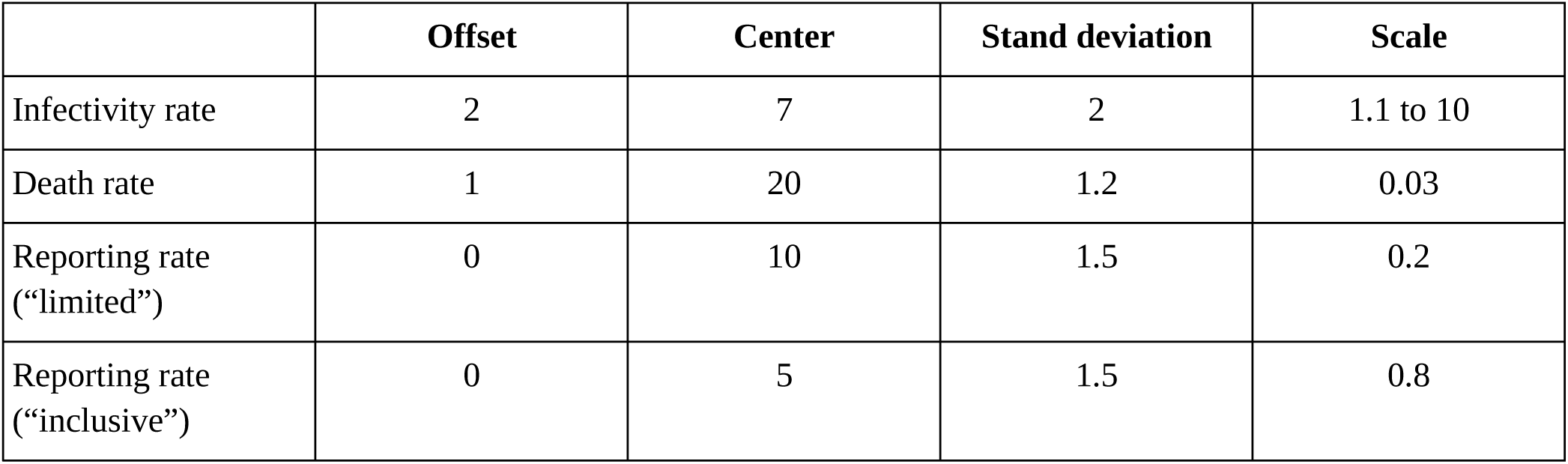
Gamma distribution^9^ parameters used for infectivity, death rates, and reporting rates. The mean of the gamma function is (center – offset), and the variance is the square of the standard deviation.

|  | Offset | Center | Stand deviation | Scale |
| --- | --- | --- | --- | --- |
| Infectivity rate | 2 | 7 | 2 | 1.1 to 10 |
| Death rate | 1 | 20 | 1.2 | 0.03 |
| Reporting rate (“limited”) | 0 | 10 | 1.5 | 0.2 |
| Reporting rate (“inclusive”) | 0 | 5 | 1.5 | 0.8 |

**Figure 1:**
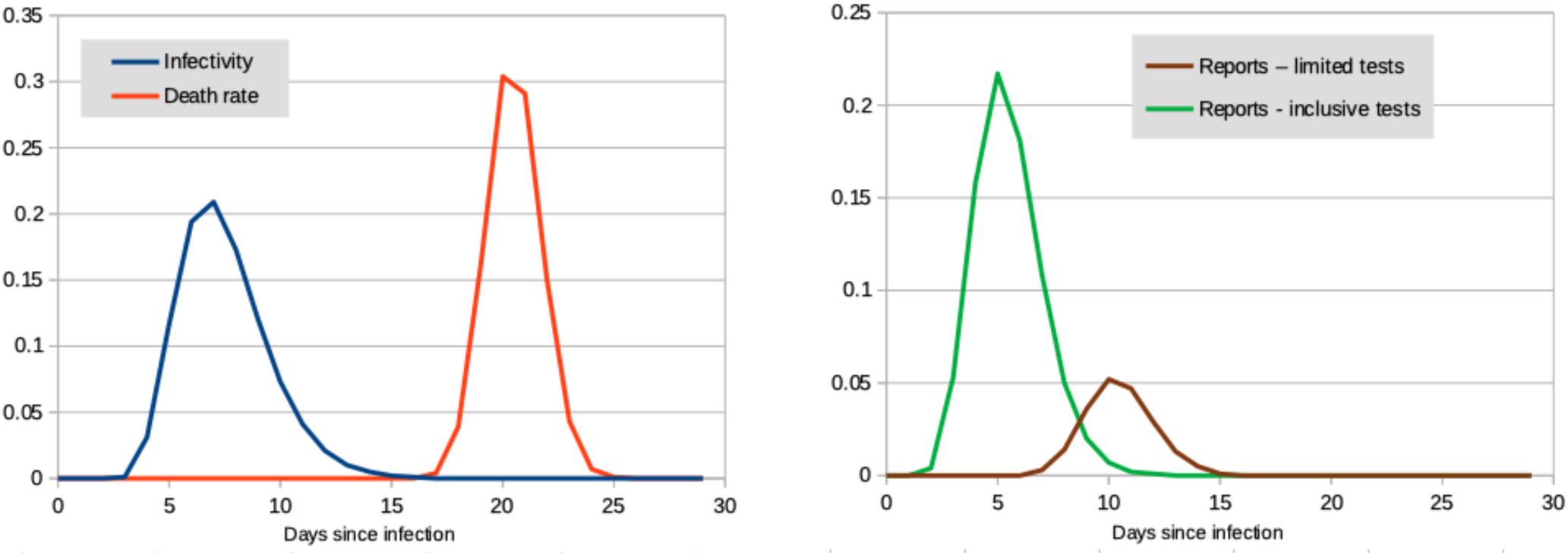
Probability functions for infection, death, and reporting

Deaths in the infected group are treated in a similar way, using an average time of 20 days between infection and death. All remaining individuals in the infected group are moved to the recovered group at the end of the 30-day time window.

For each daily iteration, we consider the daily sub-groups in the infected population as follows:

1. New infections are calculated by multiplying the number of infected individuals in the sub-group with the infectivity corresponding to the “infection age” for the group, corrected for the percentage of the original population remaining in the susceptible group.
2. Death and recovery are calculated similarly by using “infection-age” specific factors, and removed from the infected group.
3. Subsequently, all infected groups are aged by one day.
4. New infections are put in the age = 0 group and removed from the susceptible pool.

To simulate the reporting delays of confirmed cases, we used a similar approach. A day-specific reporting rate was used to reflect the variation in time between infection and the reporting of test results; daily report numbers were calculated by multiplying the counts for each infection-age groups with the age-specific reporting factor, and summing over all tracked infection ages. To account for differences in the “inclusiveness” of testing strategies, the reporting rates are scaled by a constant factor; this “reporting rate factor” reflects the percentage of COVID-19 infections that match the test criteria and are tested. For the “limited testing” scenario, we assume that 80% of the infections are asymptomatic or have only light symptoms that do not meet the minimum criteria for testing, resulting in a reporting rate factor 0.2. For “inclusive” testing, the factor was set to 0.8, based on estimates that 20% of infections are asymptomatic^8^ or would be not tested for other reasons.

Table 1 shows the parameters used to generate the per-day infectivity and death factors. Figure 1 shows a graphical representation.

## Results

### Detection rates for “limited” test policies

Figure 2 shows the predicted infections and confirmed cases over the course of an epidemic in the absence of government interventions. The maximum number of confirmed cases is about 20% of the number of infections, which reflects the reporting rate for a testing strategy that limits tests to only severe cases.

**Figure 2:**
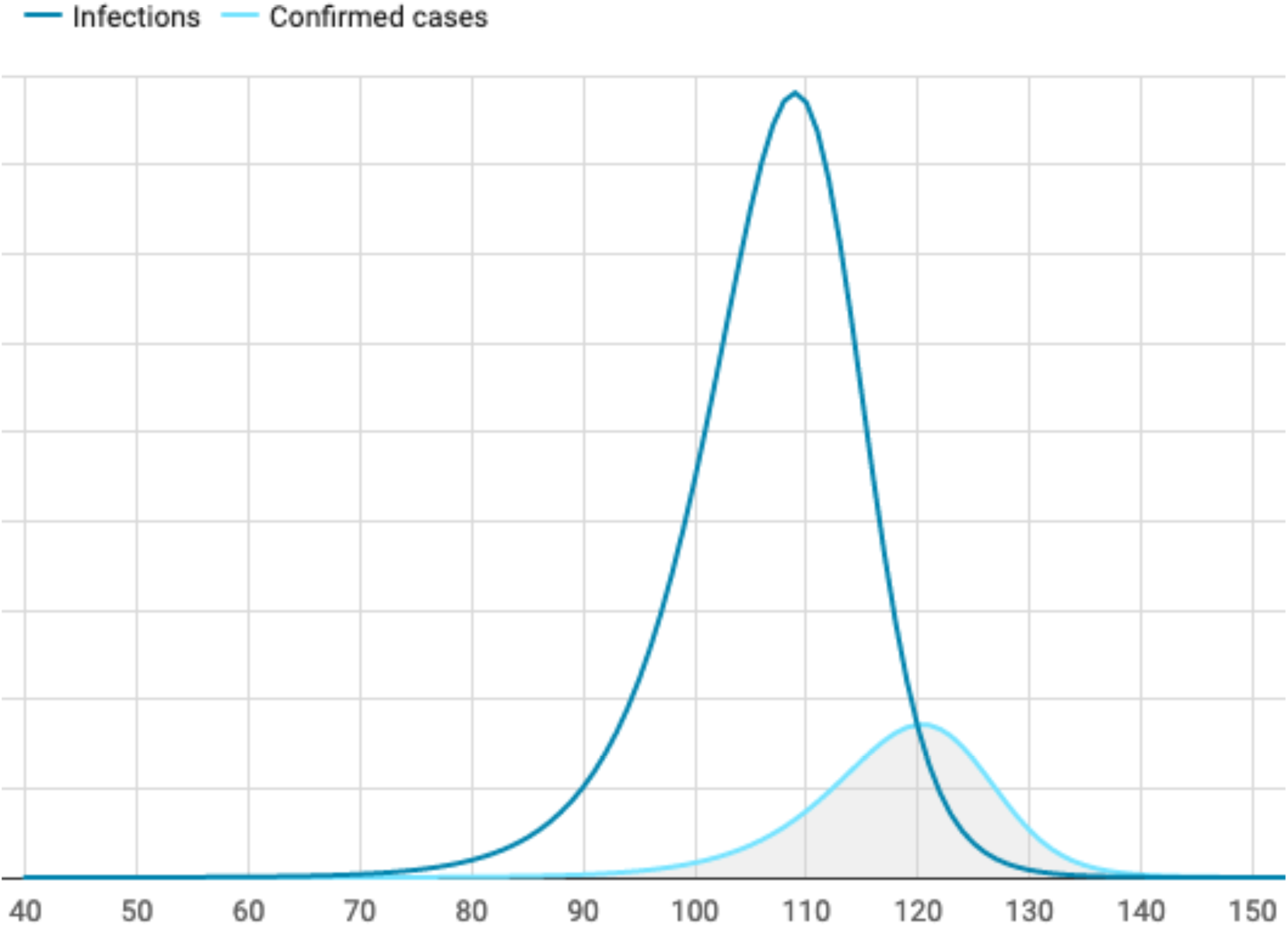
Predicted infections and confirmed cases for R^0^ = 4.0

The maximum for the confirmed cases curve is delayed by about 11 days relative to the infections curve. This reflects the following delays: (a) the incubation period of approximately 5 days (on average); (b) a period of about 2 to 3 days until symptoms become severe enough to satisfy test restriction criteria; and (c) a period of about 3 to 4 days to obtain permission for the test, perform the test, get the results, and have the results included in the officially reported numbers. While the epidemic is in the rapid growth phase, the sum of these delays creates a difference between actual infections and reported cases that is *much larger* than the testing fraction would indicate. This “under-reporting factor” depends on the growth rate of the epidemic, compared to the delay: the faster the epidemic grows, the larger the under-reporting factor will be.

To quantify the relation between the speed of the growth and the under-reporting rate, we ran the model with multiple different initial reproduction rate numbers from R^0^ = 1.1 to 10.0, and calculated the “detection rate” (defined as reported cases / actual infections per day) and the daily growth rate (defined as daily increase/previous day total) for the day where the number of reported deaths reached 1,000. The results are shown in Figure 3.

**Figure 3:**
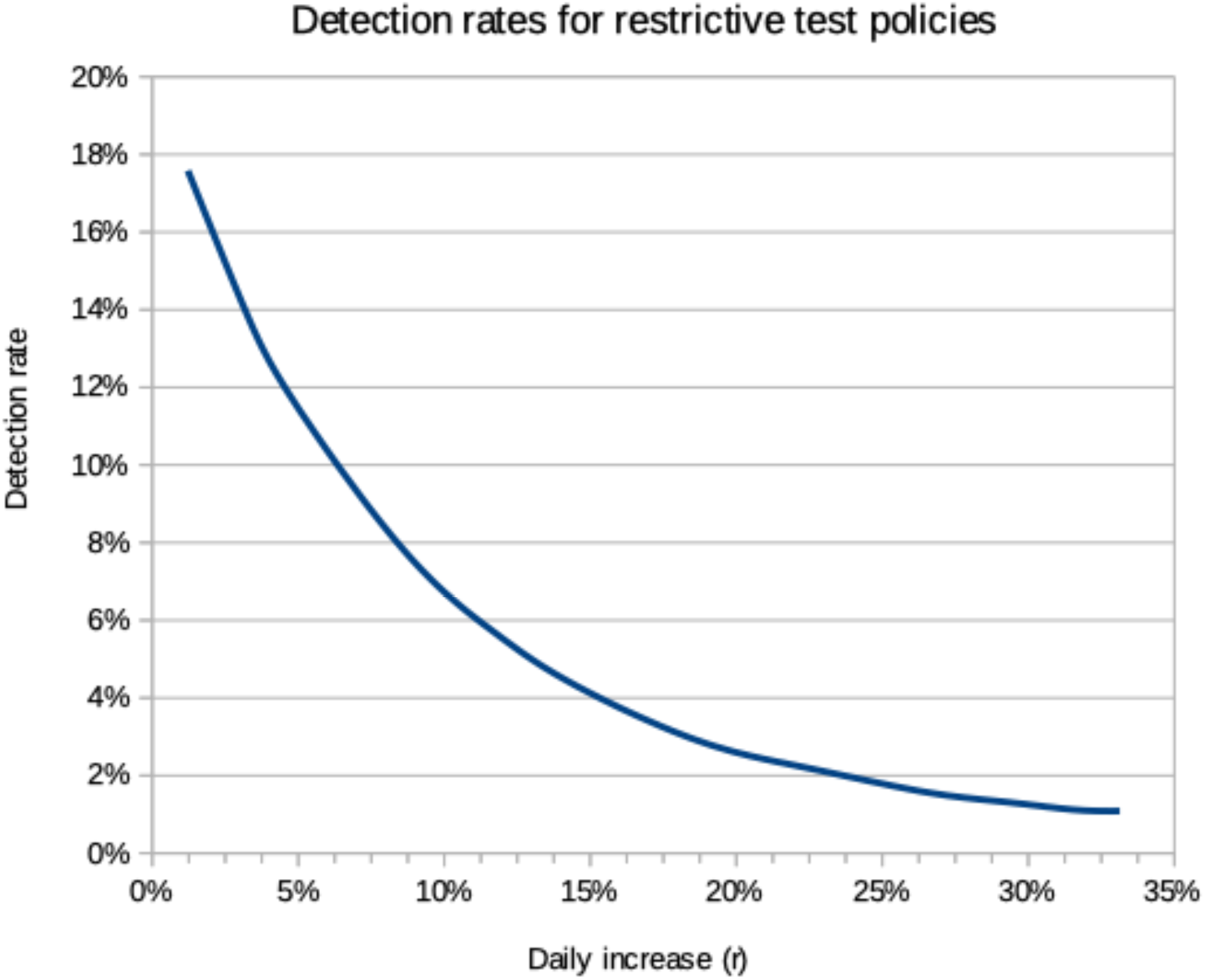
Ratio of reported confirmed cases to actual infections for different epidemic growth rates under “limited” (restrictive) testing policies

At low growth rates (R^0^ near 1.0), the observed detection rate approaches the theoretical detection rate of 20% that is dictated by the restricted test criteria, under which only about 20% all infected persons will get tested. But during faster growth phases, the detection rate drops rapidly to much lower numbers, and barely exceeds 1% when the number of new cases per day is more than 30% of the previous day’s total. But how often do COVID-19 epidemics actually reach such a growth rate?

To answer this question, we looked at the daily growth rates for the COVID-19 epidemic in six countries that reported rapid growth in March 2020. The daily growth rate was calculated as the average rate over a 7-day period; to avoid uncertainties from changes in test rates, we calculated increases from reported deaths, starting on the day the number of reported deaths exceeded 100. The results are shown in Table 2.

**Table 2:** Observed daily increases in reported deaths

| Country | Date | Increase factor | Daily increase (r) |
| --- | --- | --- | --- |
| France | 03/16/20 | 5.81 | 28.6% |
| Iran | 03/05/20 | 4.01 | 21.9% |
| Italy | 03/04/20 | 7.73 | 33.9% |
| Netherlands | 03/20/20 | 5.15 | 26.4% |
| Spain | 03/13/20 | 7.84 | 34.2% |
| USA | 03/17/20 | 5.93 | 29.0% |

All of the countries shown in Table 2 experienced very rapid growth rates. The results of this study indicates that, in conjunction with restrictive test policies, the reported number of confirmed cases was about 50 to 100-fold lower than the actual number of infections.

### Detection rates under “inclusive” test policies

In contrast to countries with restrictive test policies, some countries have “inclusive” test policies that aim to test *every* person who could potentially be infected with COVID-19. Typically, such policies are accompanied by extensive contact tracking efforts, where the aim is to identify and test every person who has been close to a person known to be infected. When such policies are implemented early in an epidemic, and sufficient testing capacities exists, they can reduce the delay between infection and confirmation of infection.

The reduction of reporting delays arises from a combination of three effects: (a) the elimination of waiting for symptoms to become severe; (b) the elimination of testing delays due to inadequate testing capacity; and (c) the testing of pre-symptomatic individuals. We ran simulations for such an “inclusive” testing regime using the following assumptions:

1. Report delays are reduced from 10 days to 5 days.
2. Testing identifies 80% of infected individuals; the remaining 20% are asymptomatic community infections, false negative tests, and similar.

The results are shown in Figure 4.

**Figure 4:**
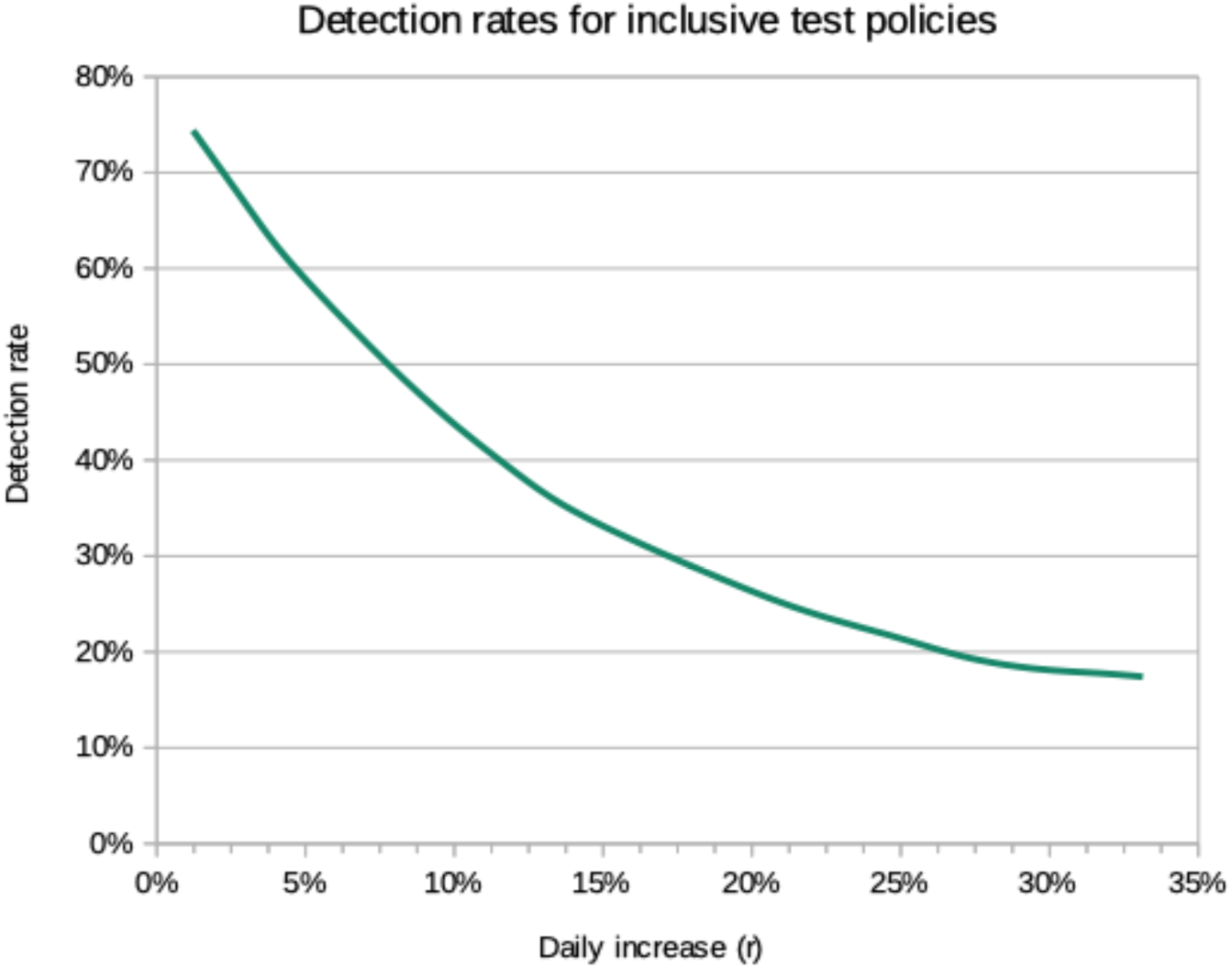
Ratio of reported confirmed cases to actual infections for different epidemic growth rates under “inclusive” testing policies and adequate testing capacity

Compared to the restrictive testing strategy, a substantially larger percentage of infections is detected. For epidemics with a daily growth of 10% or less, more than 40% of infections are detected. For very fast growing epidemics with daily growth rates above 30%, close to 20% of all infections are detected. This is about 15-times more than restrictive testing would detect. Table 3 shows a comparison of the detection rate for different epidemic growth rates.

**Table 3:** Daily increase and detection rates for different initial reproduction factors

| **R** | **Daily increase** | **Detection rate**<br>**(restrictive)** | **Detection rate**<br>**(inclusive)** |
| --- | --- | --- | --- |
| 1.1 | 1.2% | 17.58% | 74.4% |
| 1.3 | 3.2% | 14.01% | 65.9% |
| 1.5 | 5.0% | 11.52% | 59.0% |
| 2.0 | 8.7% | 7.76% | 47.4% |
| 2.5 | 11.7% | 5.70% | 39.6% |
| 3.0 | 14.3% | 4.42% | 34.3% |
| 4.0 | 18.5% | 2.96% | 28.3% |
| 5.0 | 21.9% | 2.28% | 24.2% |
| 6.0 | 24.8% | 1.82% | 21.6% |
| 7.0 | 27.2% | 1.49% | 19.4% |
| 8.0 | 29.6% | 1.29% | 18.2% |
| 9.0 | 31.6% | 1.12% | 17.8% |
| 10.0 | 33.1% | 1.09% | 17.4% |

### Detection rates after slow-down measures take effect

The data presented above are for “steady growth” scenarios – but what happens when government interventions start slowing down the epidemic growth? Two examples are shown in Figure 5 and 6. The model runs shown used a “restrictive” report delay of 10 days, but assumed a 100% test rate so that the infections and confirmed cases can be compared more easily.

**Figure 5:**
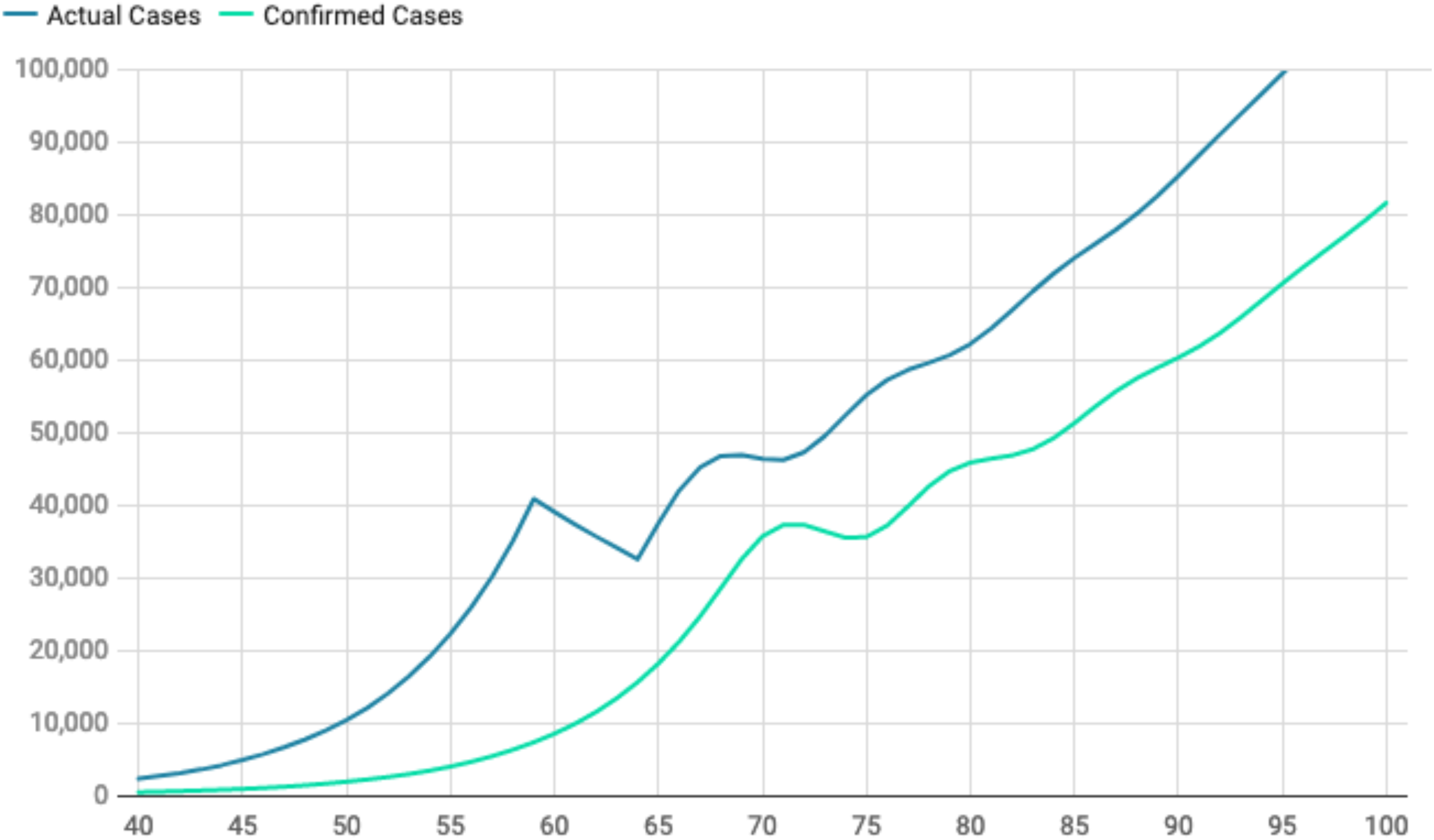
Predicted case and confirmation numbers after government interventions reduce R^0^ from 3.5 to 1.3; adaption of measures is gradual over 6 days. The confirmed cases curve assumes that 100% of all infections are tested. This is only done to enable a direct comparison; actual test rates in most countries are significantly lower.

**Figure 6:**
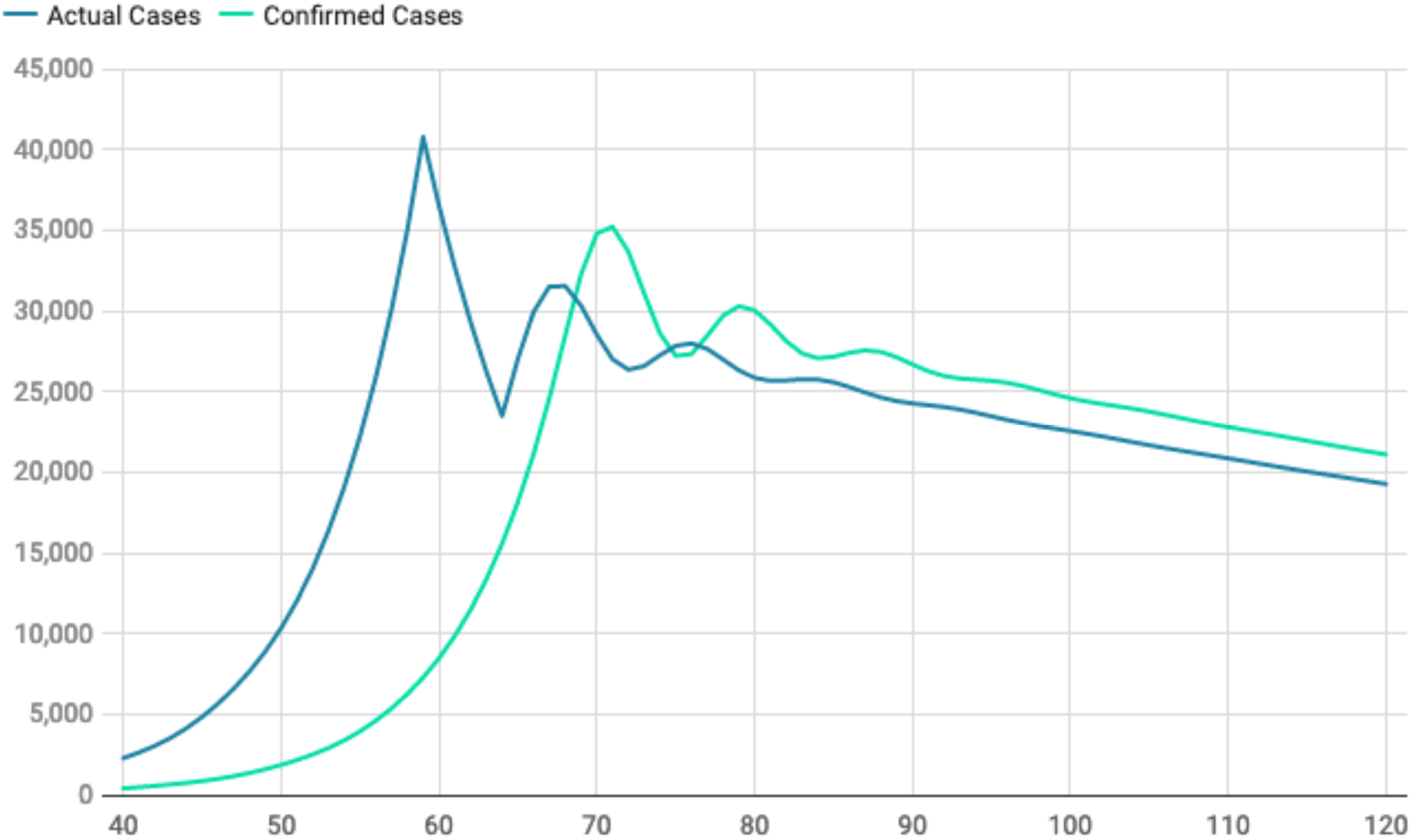
Predicted case and confirmation numbers after government interventions reduce R^0^ from 3.5 to 0.94; adaption of measures is gradual over 6 days. The confirmed cases curve assumes that 100% of all infections are tested. This is only done to enable a direct comparison; actual test rates in most countries are significantly lower.

Figure 5 shows the prediction for interventions that are only partially effective, dropping the initial reproduction number from 3.5 to 1.3. The drop is modeled to take place over a period of 6 days, reflecting gradual adaptation by the population or by different regions. After interventions begin taking effect on day 59, new daily infections start to drop, but the number of confirmed cases keeps going up for another 10 days, reflecting reporting delays. The sharp drop in the actual infections is smoothed out into a sinusoidal shape in the confirmed cases; this is because infections from a given day will not all be reported on the same day due to differences in symptom onset times and differences in reporting delays.

The under-reporting factor changes as the growth of the epidemic changes, dropping from higher values during the initial rapid growth phase to lower values during the slower growth phase. This simply reflects that relatively fewer new infections happen during the time between initial infection and actual confirmation of cases in the slower growing phases.

Figure 6 shows the relation between actual infections and confirmed case numbers for a scenario where more effective government interventions reduce the reproductive rate to 0.94, causing a steady drop in new daily infections. This causes a significantly sharper drop in new infections during the adaptation phase, which is also reflected in the number of confirmed case. During the slowly declining phase of the epidemic, the reporting delay causes the number of confirmed cases to be slightly higher than the actual number of new infections. Note, however, that the figure assumes *all* infected persons will be tested; under restrictive testing strategies, the actual number of tested persons would likely be much lower, so that that the confirmed cases will still under-report actual infections significantly.

## Discussion

In many countries, the response to the COVID-19 epidemic followed a similar pattern: an initial phase where the threat was ignored or downplayed, and no actions were taken; a second phase with limited actions, for example the prohibitions of large gatherings, partial border closings, and *suggestions* to practice social distancing; and, eventually, a third phase marked by multiple, increasingly stricter interventions, and enhanced enforcement of regulations.

How fast countries switched to more advanced response phases was largely determined by the *perceived threat level*. Several countries and regions that had been hit hardest by the SARS and MERS epidemics quickly jumped to phase 2 and 3 measures, and were generally successful in containing the epidemic. As of April 10, 2020, Taiwan, Singapore, and Hong Kong each had fewer than 10 deaths from COVID-19, despite seeing early COVID-19 cases after the initial cases in China.

In stark contrast, many other countries had much longer delays before starting intense interventions; these delays led to more than 1,000 COVID-19 deaths in 13 countries. In these countries, the number of reported COVID-19 cases in each country was the main determinant in the perceived threat level. Generally, the number of confirmed cases is taken *at face value*, and it had to rise to a high level before governments felt strict interventions were justified, and before the population was willing to adhere to imposed restrictions.

This study shows that **the number of confirmed cases understates actual infections during the rapid growth phase of a COVID-19 epidemic by a factor of as much as 50 to 100**. This rather large factor arises primarily from the rapid growth of the epidemic and the delays between infection, symptom onset, testing, and reporting of results. This is compounded by testing policies that restrict access to testing in most countries, and test delays caused by insufficient testing capacities.

While we used a computer model to illustrate our findings and get numerical estimates, it should be noted that the results are not specific to the model employed. Very similar results can be obtained by using observed daily growth rates of an epidemic, and existing knowledge about the delay between infection and the onset of symptoms that meet the existing test criteria. The actual numbers may vary slightly, but the overall conclusions will remain the same.

It should also be noted that the under-reporting factor of 50 to 100 is *not* an upper bound, since we did not consider other factors like, for example, false-negative test rates.^10,11^

To obtain an accurate picture of the state of a rapidly growing epidemic, *early adequate testing capacity is essential*. Insufficient test capacity can cause delays and underestimates through multiple factors. These include test backlogs, like reported delays of up to 10 days to get test results in California^12^; additional delays by requiring symptoms to be severe; and capacity-mandated limitations to who can be tested. Our results show that these factors alone can cause a 15-fold under-reporting of infections, compared to an “inclusive” testing strategy. To reduce substantial under-estimating of the severity of the COVID-19 epidemic, it is essential to (a) include as many potentially infected persons, and (b) to have adequate test capacity with fast turnaround times.

The results of this study also show that under-reporting factors go down when government interventions take effect and reduce the growth rate of an epidemic. However, underreporting factors can still be substantial even at slower growth rates, especially if testing is limited to a subset of infected persons. Often, local testing bottlenecks can create this situation even if the stated policies are not overly restrictive.

While the growth rate of the epidemic appears to be slowing down in many countries, which should reduce the under-reporting of infections to some extend, regional hot spots of growth remain; in these regions, under-reporting is likely to still be a larger problem. Increased test capacities, fast turnaround times, and inclusive testing policies will also be essential when attempts are made to gradually relax restrictions; otherwise, test and reporting delays could cause a substantial *undetected* growth if measures are relaxed too much.

## Data Availability

Data used for this study were downloaded from the public repository at https://github.com/CSSEGISandData/COVID-19/

